# Metabolite patterns of patients with peripheral arterial disease in response to exercise

**DOI:** 10.1101/2021.07.24.21261067

**Authors:** Tiffany R. Bellomo, Noah L. Tsao, Hillary Johnston-Cox, Kamil Borkowski, Gabrielle Shakt, Renae Judy, Jonni S. Moore, Sarah J. Ractcliffe, Oliver Fiehn, Emile Mohler, John W. Newman, Scott M. Damrauer

## Abstract

Supervised exercise therapy (SET) is an effective intervention for symptomatic peripheral artery disease. Its effect on metabolism, measured by the circulating metabolome is not well understood. Participants underwent the Gardner graded treadmill test before and after SET and blood was sampled before and after each treadmill test. We tested the average association of metabolite levels with timing of blood draws. We used five models to identify metabolites or changes in metabolites at specific time points that associate with treadmill test performance or inter-individual variability in functional performance after SET. When analyzing individual time points, high levels of anandamide (AEA) before any exercise interventions were associated with shorter, or worse, walking time. Increased arachidonic acid (AA) and decreased levels of AA precursors (dihomo-γ-linolenic acid and diacylglycerol) before any exercise was associated with shorter walking times. Participants who tolerated large increases in AA during acute exercise had longer, or better, walking times before and after SET. We identified two pathways of relevance to individual response to SET: AEA synthesis may increase the activity at endocannabinoid receptors, resulting in worse treadmill test performance. SET may help train patients withstand higher levels of AA and inflammatory signaling, resulting in longer walking times.

## INTRODUCTION

Peripheral arterial disease (PAD) is progressive occlusive atherosclerotic disease of the aorta and lower extremities that affects more than 200 million people worldwide (1). Insufficient blood flow to the lower extremities is quantified in part by the ankle brachial index (ABI) and leads to ischemia driven symptoms of PAD, ranging from leg pain with ambulation to ulceration and tissue loss. Claudication is a classic symptom of early PAD defined as reproducible discomfort or fatigue in the muscles of lower extremities that is brought on with ambulation (2). The physical activity limitation of patients with PAD results in functional impairment, loss of mobility, and decreased quality of life (3, 4).

Treatment options available for PAD include a combination of supervised exercise therapy (SET), medications, and revascularization (5–9). The efficacy of SET in patients with PAD dates back to 1966 when Larsen et. al demonstrated that 6 months of intermittent walking therapy improved walking time to onset of discomfort and peak walking time (PWT), which represents claudication limited exercise tolerance (10). Over the last 3 decades, there are have been multiple clinical trials and meta-analyses that have added to our understanding of the benefits of SET with and without revascularization versus optimized medical therapy (11, 12, 21, 13–20). However, there has not been consistent evidence for substantial benefit using one intervention strategy over another due to inter-individual variability in response to therapies (22–24).

The pathophysiology underlying not only functional decline, but also the effects of exercise observed in patients with PAD are complex and poorly understood (25). It is thought that both structural as well as metabolic disarray in calf skeletal muscle affects contractile performance and contributes to walking impairment, pain, and functional decline. Although SET does not affect plaque morphology or axial artery blood flow, there have been several proposed mechanisms of action in PAD: increased calf blood flow via the microvasculature, improved endothelial function and subsequent improved vasodilatation, reduced inflammation, improvements in muscle structure, strength and endurance, vascular angiogenesis, improved mitochondrial function, and skeletal muscle metabolism through lipid-based signaling (5, 26–32). Despite the fact that metabolomics have been used to investigate some of these proposed mechanisms in the context of PAD progression (33, 34), metabolomics have not been leveraged to examine the response to exercise training in patients with PAD. Investigating the effects of SET on the metabolome may provide insight into underlying mechanisms and pathophysiology of exercise training in the context of PAD and identify circulating biomarkers reflective of the inter-individual variability in the response to exercise therapy.

In the current study, metabolomic and lipidomic techniques were utilized to measure the effects of SET on primary metabolites, complex lipids, and lipid mediators. These effects were then correlated with individual, subject-level measures of the response to SET among subjects with PAD. We identified metabolites that are associated with the inter-individual variability in the response of subjects with PAD to SET. This insight will enhance our understanding of the pathophysiology of lower extremity symptoms in PAD and the mechanism by which SET improves walking intolerance.

## RESULTS

### Trial Design and Demographic Characteristics

This study was conducted on a subset of individuals enrolled in a randomized trial of SET (35, 36). In the overall trial there were no significant differences between the characteristics of participants in the control and treatment cohorts: participants were a median 67 years old [IQR 60.3-73.0] with a body mass index (BMI) of 28 [IQR 25.1-30.6] and 64% were male (Supplemental Table 1). Although pre-training ABI was significantly different between the control and treatment cohorts (median [IQR]; control = 0.65 [0.59-0.72], treatment = 0.68 [0.61-0.86, p = 0.045, these differences are not clinically significant, and post-training ABI was not significantly different between the cohorts.

### Metabolite Data Cohort

To identify metabolites associated with the inter-individual variability in the response of subjects with PAD to SET, 40 participants who were randomized to the exercise program arm and completed at least 80% of prescribed sessions were chosen for metabolite analysis. These 40 participants were a median of 65 years old [IQR 60.8-69.3] with a BMI of 28 [IQR 25.8-30.1] and 75% were male (Table 1).

**Table 1.**
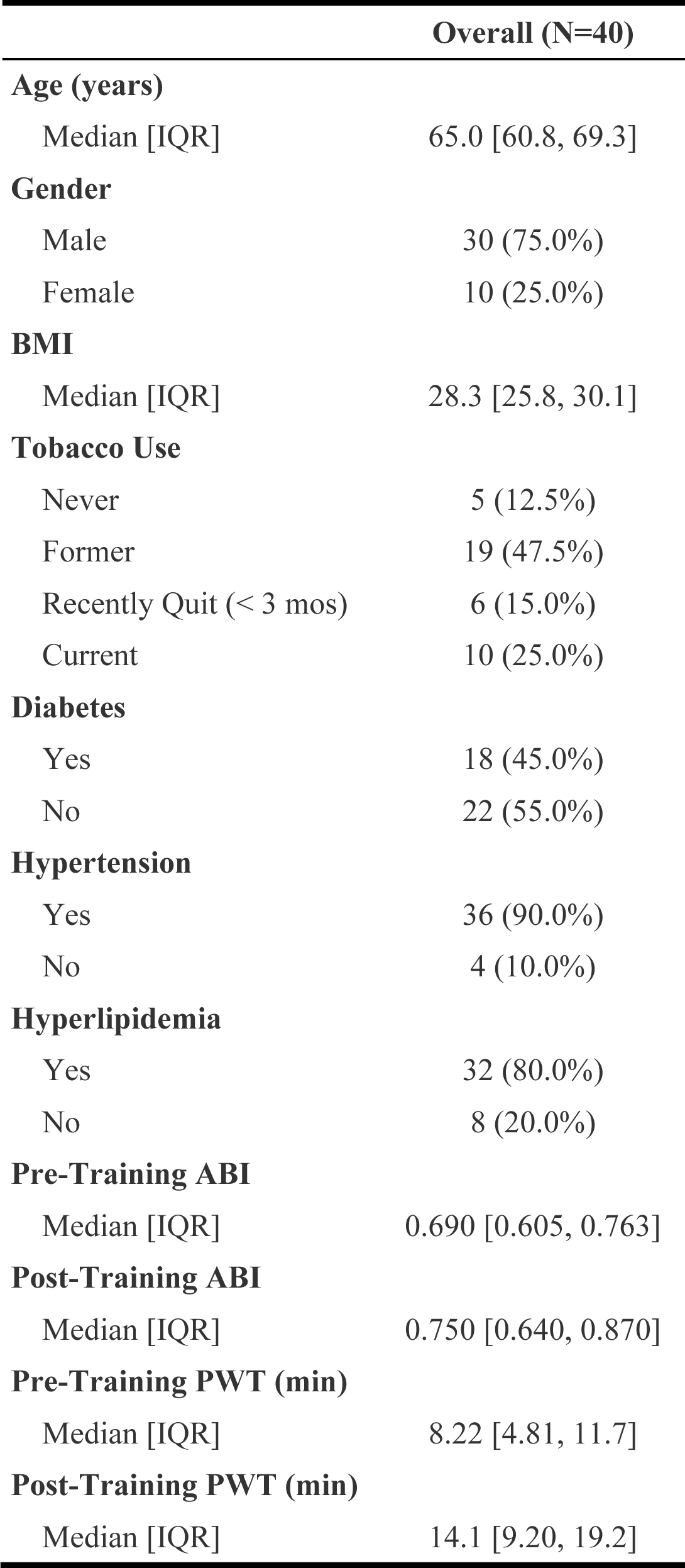
Demographic characteristics of participants (n = 40) who had plasma samples drawn before and after treadmill testing.

|  | Overall (N=40) |
| --- | --- |
| **Age (years)** |  |
| Median [IQR] | 65.0 [60.8, 69.3] |
| **Gender** |  |
| Male | 30 (75.0%) |
| Female | 10 (25.0%) |
| **BMI** |  |
| Median [IQR] | 28.3 [25.8, 30.1] |
| **Tobacco Use** |  |
| Never | 5 (12.5%) |
| Former | 19 (47.5%) |
| Recently Quit (< 3 mos) | 6 (15.0%) |
| Current | 10 (25.0%) |
| **Diabetes** |  |
| Yes | 18 (45.0%) |
| No | 22 (55.0%) |
| **Hypertension** |  |
| Yes | 36 (90.0%) |
| No | 4 (10.0%) |
| **Hyperlipidemia** |  |
| Yes | 32 (80.0%) |
| No | 8 (20.0%) |
| **Pre-Training ABI** |  |
| Median [IQR] | 0.690 [0.605, 0.763] |
| **Post-Training ABI** |  |
| Median [IQR] | 0.750 [0.640, 0.870] |
| **Pre-Training PWT (min)** |  |
| Median [IQR] | 8.22 [4.81, 11.7] |
| **Post-Training PWT (min)** |  |
| Median [IQR] | 14.1 [9.20, 19.2] |

The median peak walking times before and after SET were 8.2 [IQR 4.81-11.7] and 15.0 [IQR 9.20-19.2] minutes respectively. No demographic factors tested (age, sex, BMI, smoking status, diabetes, heart attack or stroke, hypertension, hyperlipidemia, minimum ABI, maximum ABI) associated with baseline PWT. Baseline maximum ABI (β=1.49 unit change in ABI per min, p = 0.008 [95% CI = 0.44 -2.54], FDR = 0.18) and minimum ABI (β=1.41 unit change in ABI per min, p = 0.02 [95% CI = 0.26 – 2.55], FDR = 0.23) had nominally significant positive associations with post SET-PWT. These associations did not pass multiple testing correction. The median change in PWT was 5.0 min [IQR 0.8-9.3]. Age had a nominally significant negative (β=-0.05 min per year, p=0.04, [95% CI = -0.10 - -0.01], FDR = 0.15) association with PWT, however, these associations also did not pass correction for multiple testing.

Metabolite levels were assessed before and after Gardner graded treadmill tests that took place before and after a 12-week SET program (Figure 1) (37). Plasma samples were analyzed for 3 different metabolite panels: primary metabolites measured by GC-TOF, complex lipids measured by LC-TOF, and lipid mediators measured by UPLC QTRAP. To reduce multiple testing and focus on identifiable biological mechanisms, only peaks corresponding to known metabolites and metabolites with complete data for all samples were considered for analysis (Supplemental Figure 1). In the primary metabolite panel, 364 metabolites were measured, 122 peaks corresponded to known metabolites, and no metabolites were excluded for incompleteness. In the complex lipids panel, 2569 metabolites were measured, 327 peaks corresponded to known metabolites, and 21 metabolites were excluded for incompleteness. In the lipid mediators panel, 66 metabolites were measured, all of which corresponded to known metabolites, and 2 metabolites were excluded for incompleteness. All metabolites used for analyses were annotated with RefMet (38) compound classifications in anticipation of performing hypergeometric enrichment analysis to identify enriched classes of metabolites in each analysis. For enrichment analysis, nominal significance was defined as p < 0.05 and experiment wide significance was defined as a Benjamini-Hochberg false discovery rate (FDR) < 0.05.

**Figure 1.**
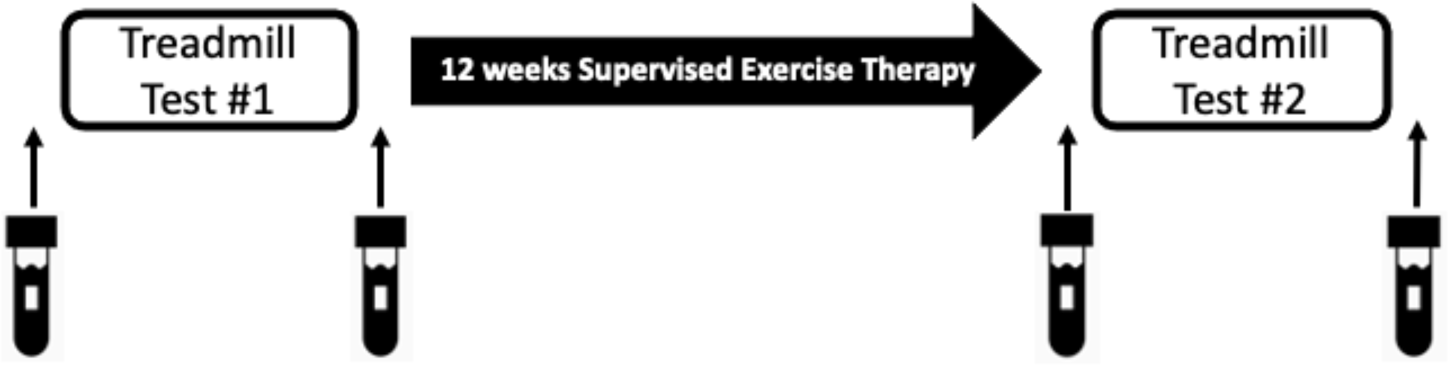
Experimental design for each participant (n = 40). Participants underwent a Gardner graded treadmill test before and after 12 weeks of supervised exercise therapy. A plasma sample is drawn before and after each Gardner graded treadmill test for metabolomic analysis.

### Diabetes and BMI were associated with carbohydrate metabolites

Baseline metabolite levels (blood draw before the first treadmill test) were tested for an association with the following outcomes, controlling for age and sex: minimum ABI, maximum ABI, smoking status, diabetes, and BMI. There were numerous nominally significant (p < 0.05) metabolites associated with these demographic factors (Supplemental Table 2). Diabetes status was significantly associated with glucose after multiple testing correction (β=0.64, p<0.001, OR=1.90 [95% CI: 1.39-2.59], FDR = 0.04) (Supplemental Table 3). BMI was also significantly associated with glucose (β=0.30, p<0.001, OR=1.36 [95% CI: 1.15-1.60], FDR = 0.049) and mannose (β=0.33,p<0.001, OR=1.40 [95% CI: 1.17-1.70], FDR = 0.049) levels after multiple testing correction, verifying previously known associations (39).

### SET had minimal average effect on the metabolome

We first sought understand the average impact of acute exercise and SET on the metabolome. We used repeated measures, mixed-effects, generalized linear models to test the association between acute exercise (treadmill test), SET, and their interaction with metabolite levels measured at all 4 blood draws (Supplemental Figure 2; Supplemental Table 4). In these analyses, the levels of 12 metabolites were found to associate with acute exercise (Supplemental Table 5); 11 of these 12 metabolites were in the lipid mediators data set. We did not detect any metabolite levels that associated with either SET or interaction of SET and treadmill test, suggesting that on average, there was minimal effect of SET on the metabolome.

Enrichment analysis performed using a hypergeometric test on the 11 lipid mediators (Supplemental Table 6) determined classes of lipid mediators that were over-represented: organonitrogen compounds were enriched at nominal significance (p=0.03, FDR = 0.06) and fatty amides were enriched at experiment wide significance (FDR = 5E-4) (Supplemental Figure 4). These have both been previously shown to change as a result of acute exercise and effect downstream endocannabinoid signaling in response to exercise challenges (40). In addition, Acyl ethanolamide (AcylEA) metabolites, including anandamide (AEA), N-oleoylethanolamide (OEA), linoleoyl ethanolamide, adrenoyl-ethanolamide (C22:5n6-ethanolamide; DEA) and cervonoyl ethanolamide (C22:6n3-ethanolamide; DHEA) were all on average negatively associated with acute exercise, meaning average levels of AcylEAs in plasma decreased after acute exercise. Although OEA has been previously reported to increase with exercise and hypothesized to signal exertion and fatigue, this has only been demonstrated to occur locally in muscle tissue (41).

### Metabolite levels before acute exercise associate with performance during acute exercise

Next, we sought to understand the association between baseline metabolite levels with absolute performance on the treadmill test, as measured by peak walking time (PWT) (Figure 1). To do this, we used five different models (general linear models [GLM], least absolute shrinkage and selection operator regression [LASSO], elastic net, random forest [RF], and support vector regression [SVR]) to identify metabolites or changes in metabolites at specific time points that associated with treadmill test performance (Figure 2). Metabolites of significant interest were defined as having achieved nominal significance (p < 0.05) in 2 or more models or experiment wide significance in one model with a Benjamini-Hochberg FDR < 0.05. We then performed enrichment testing to identify classes of metabolites of significant interest that associate with exercise (Figure 3).

**Figure 2.**
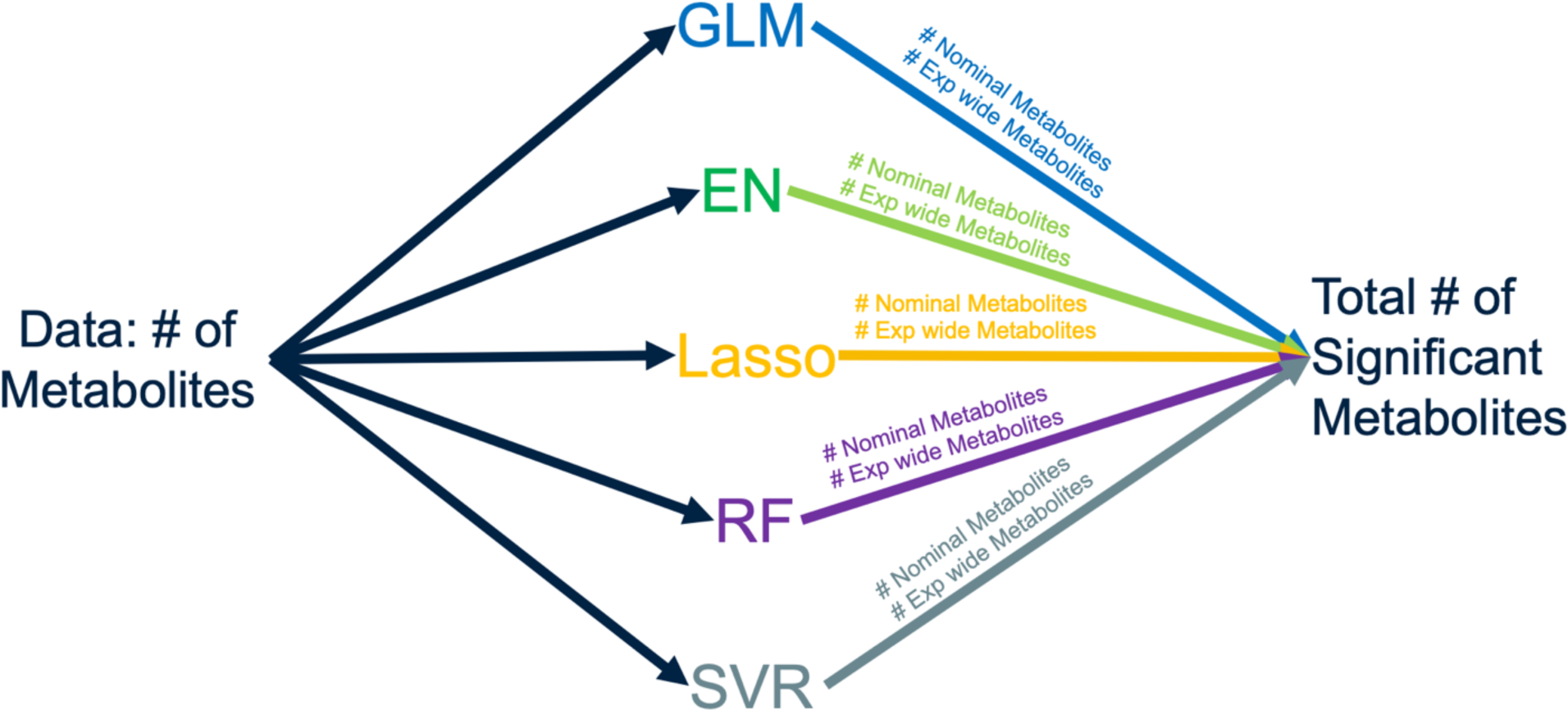
Scheme for defining metabolites of significant interest. Metabolites were analyzed in relationship to pre-SET natural logarithm transformed peak walking time and the standardized measure of individual improvement in peak walking time for each of the three data sets using 5 models: general linear model (GLM), elastic net (EN), lasso, random forest (RF), and support vector machine (SVR). Metabolites of significant interest were classified as achieving nominal significance on two or more models or experiment wide significance on one model. Multiple testing correction threshold was set at a false discovery rate of 0.05 for each model and for each of the three data sets.

**Figure 3.**
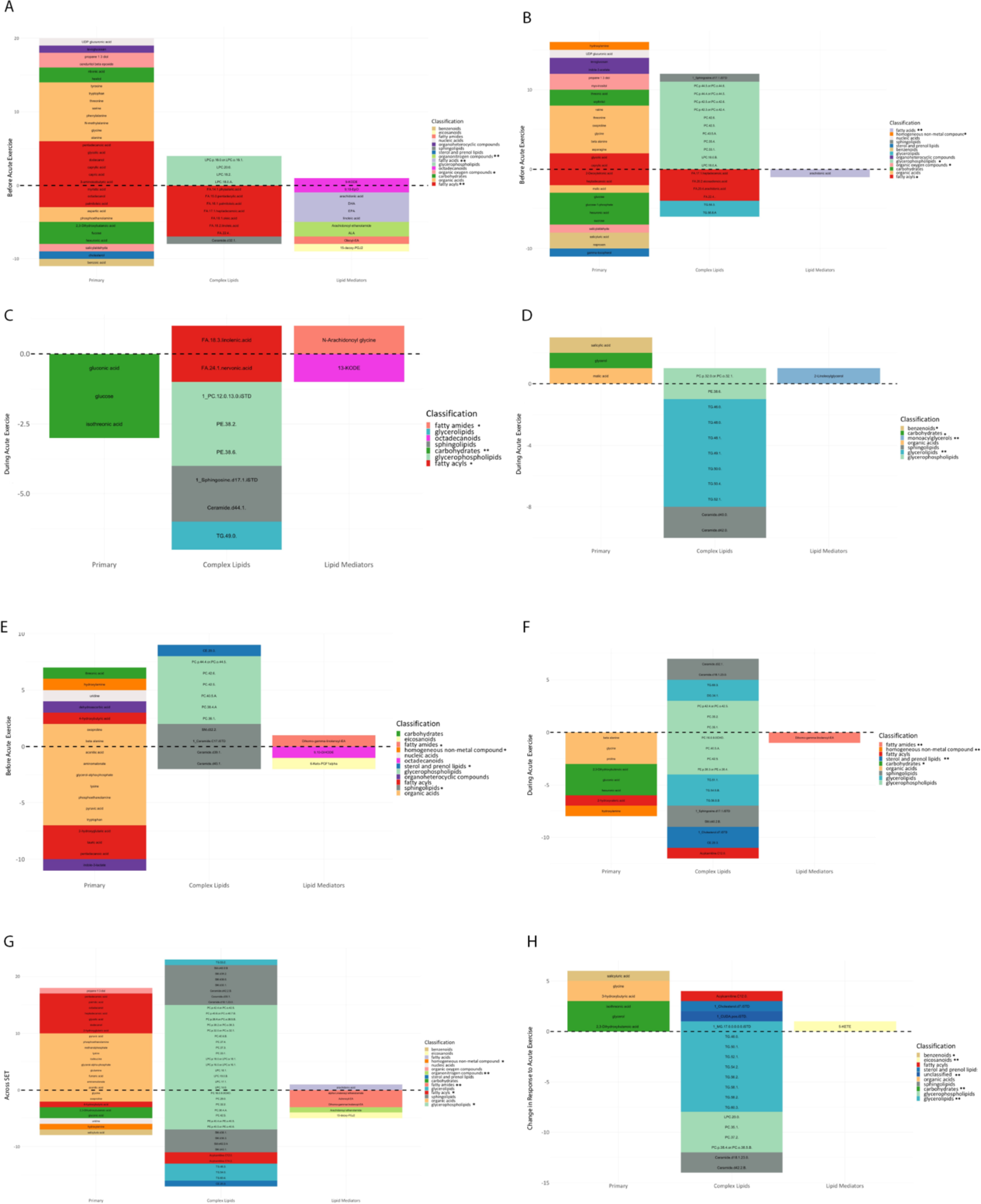
Metabolites or changes in metabolites at specific time points that associate with either treadmill test performance or inter-individual variability in functional performance after SET. Five different models (general linear model, least absolute shrinkage and selection operator regression, elastic net, random forest, and support vector machines) were used to analyze primary, complex lipid, and lipid mediator metabolite data sets. Metabolites of significant interest were defined as having achieved nominal significance (p<0.05) in 2 or more models or experiment wide significance in one model with a Benjamini-Hochberg FDR<0.05. Only direction of association is displayed given that 5 models were used in combination to define metabolites of significant interest. Enrichment analysis was performed to identify enriched classes of metabolites shown in the figure legend for each analysis. * indicates the RefMet classification was enriched at nominal significance. ** indicates the RefMet classification was enriched at experiment wide significance. A) Resting baseline metabolite levels prior to the first treadmill test associated with performance on the first treadmill test. B) Resting metabolite levels drawn prior to the second treadmill test associated with performance on the second treadmill test. C) Changes in metabolite levels as a result of acute exercise associated with performance on the first treadmill test. D) Changes in metabolite levels as a result of acute exercise associated with performance on the second treadmill test. E) Resting baseline metabolite levels prior to the first treadmill test associated with the standardized measure of individual improvement in peak walking time. F) Changes in metabolite levels as a result of acute exercise associated with the standardized measure of individual improvement in peak walking time. G) Resting metabolite levels drawn before each treadmill test associated with the standardized measure of individual improvement in peak walking time. H) Change in the metabolome’s response to acute exercise before and after SET associated with the standardized measure of individual improvement in peak walking time.

We conducted the primary analysis using data from the study visit before the first treadmill test and before participants underwent SET. In total, nominal significance in one model was achieved by 65 metabolites, nominal significance in two or more models was achieved by 49 metabolites, and experiment wide significance in one or more models was achieved by 4 metabolites, resulting in a total of 53 metabolites of significant interest (Supplemental Table 7). In the primary metabolites data set, the set of organic oxygen compounds was a nominally enriched group (p=0.01) and largely negatively associated with PWT (Supplemental Table 8, Supplemental Figure 4). In the complex lipid data set, the fatty acyl group was experiment wide enriched (FDR = 2E-06) and negatively associated with PWT (Supplemental Table 8, Supplemental Figure 4). In the complex lipid data set, organonitrogen compounds and fatty acids were enriched at an experiment wide level and on average negatively associated with performance on the first treadmill test (Supplemental Figure 4). This means that high levels of the organonitrogen metabolite AEA, i.e. N-arachidonoylethanolamide, and fatty acid metabolites before any exercise challenge were associated with low PWT or worse performance on treadmill test 1.

A secondary analysis tested the association of resting metabolite levels drawn prior to the second treadmill test at the time of the second study visit, after the participants had undergone 12 weeks of SET. In total, nominal significance in one model was achieved by 91 metabolites, nominal significance in two or more models was achieved by 45 metabolites, and experiment wide significance in one or more models was achieved by one metabolite, resulting in a total of 46 metabolites of significant interest (Supplemental Table 9). Our secondary analysis replicated 38 metabolites and one class of enrichment found in our primary analysis. We furthermore identified 75 additional metabolite associations. In the primary metabolites data set, the previous results for organic oxygen compounds before any exercise were replicated in this analysis after SET: organic oxygen compounds was a nominally enriched group (p=8E-03, FDR = 0.08) and salicylaldehyde was again negatively associated with PWT (Supplemental Table 10, Supplemental Figure 5). Higher levels of salicylaldehyde before acute exercise was associated with worse performance on treadmill tests, even after SET. Levels of salicylaldehyde have been previously shown to increase after acute exercise in 40 sedentary women, indicating that benzoic acid metabolism and NADPH-dependent reactions that generate salicylaldehyde as a byproduct are important mechanisms underlying acute exercise changes in the metabolome (42).

Homogenous non-metal compounds were also nominally enriched (p=0.05, FDR = 0.27) and positively associated with PWT. In the complex lipid data set, the results for fatty acyls were replicated at a nominal level; fatty acyls were nominally enriched (p=0.02, FDR = 0.07) and negatively associated with PWT (Supplemental Table 10, Supplemental Figure 5). Although no results from the lipid mediator data set were replicated from the previous analysis, the fatty acid group was enriched at an experiment wide level (p<1E-10, FDR<1E-10) and each individual metabolite was negatively associated with PWT (Supplemental Table 10, Supplemental Figure 5). Arachidonic acid (AA), a metabolite of significant interest in both the fatty acyl and fatty acids groups, has been studied as an important inflammatory mediator that undergoes dynamic changes in response to exercise that may be augmented in unhealthy human subjects (43, 44). Although it is tempting to attribute the differences in the primary and secondary analysis described above to the result of SET, this experiment was not designed to test this hypothesis.

### Dynamic changes in metabolite levels from acute exercise associate with performance during acute exercise

To better understand the relationship between acute exercise and its effects on the metabolome, we tested the association of changes in metabolite levels as a result of exercise with absolute performance on the first treadmill test. The primary analysis was conducted at the first study visit prior to participants undergoing SET. In total, nominal significance in one model was achieved by 127 metabolites, nominal significance in two or more models was achieved by 15 metabolites, and none of the metabolites achieved experiment wide significance in one or more models, resulting in a total of 15 metabolites of significant interest (Supplemental Table 11). Carbohydrates were enriched at an experiment wide level (FDR < 1E-10) in this model and negatively associated with PWT (Supplemental Table 12, Supplemental Figure 6). Therefore, large increase in carbohydrates after acute exercise were associated with low PWT or worse performance on the first treadmill test, demonstrating a previously known association (45).

Isothreonic acid (i.e. D-threonic acid), which is an uncommon carbohydrate, was negatively associated with PWT (Supplemental Table 12, Supplemental Figure 6). Therefore, large increases in isothreonic acid during acute exercise were associated with worse performance on a treadmill test. Although not much is known, isothreonic acid has been studied in healthy rats and shown to increase over acute exercise, which may reflect changes in carbohydrate metabolism in otherwise health subjects (46). Similarly, acute exercise in sedentary, obese insulin resistant women lead to an increase in carbohydrate metabolism reflected in elevations of fructose and maltose, that were correlated to changes in threonic and isothreonic acid, further supporting this contention (42) Fatty amides (FDR = 0.03) and fatty acyls (p = 0.03, FDR = 0.1) were again enriched and large increases in fatty amides after acute exercise were associated with better performance on the first treadmill test (Supplemental Table 12, Supplemental Figure 6). Notably, fatty acyls such as alpha-linolenic acid have been shown to decrease the plasma concentration of the inflammatory cytokine interleukin (IL)-6 in humans (47) and impair arterial thrombus formation in mice (48), although the mechanisms of action remain unclear.

A secondary analysis was conducted using metabolites measured at the second study visit after participants had undergone 12 weeks of SET. Nominal significance in one model was achieved by 110 metabolites, nominal significance in two or more models was achieved by 15 metabolites, and experiment wide significance in one or more models was achieved by one metabolite, resulting in a total of 16 metabolites of significant interest (Supplemental Table 13). Our secondary analysis replicated 2 metabolites, but no classes of enrichment found in our primary analysis. We identified 31 additional metabolite associations. In the primary metabolites data set, the previous results for benzenoids before SET were replicated in this experiment after SET at an experiment wide level (FDR = 0.02) (Supplemental Table 14, Supplemental Figure 7). Both glycerolipids (FDR = 4E-4) and monoacylglycerols (FDR < 1E-10) were enriched at an experiment wide level and negatively associated with PWT on treadmill test 2. Therefore, large increases in triglyceride levels during acute exercise were associated with worse performance on a treadmill test after SET. It is notable that acute resistance, but not endurance, exercise has been associated with an increase in triglyceride clearance in healthy men (49). Similarly, exercise has been shown to increase muscle lipoprotein lipase activity, promoting very low density lipoprotein clearance when intramuscular triglycerides are depleted (50). Since skeletal muscle triglyceride synthesis capacity has been shown to increase after acute exercise, higher intramuscular triglyceride reserves may reduce muscle LPL activity during exercise, leading to the observed elevation in plasma triglycerides (51, 52). Although it is tempting to attribute the differences in the primary and secondary analysis described above to the result of SET, this experiment was not designed to test this hypothesis.

### Metabolite levels before SET associate with the standardized measure of individual improvement in PWT

Effects of SET on metabolite levels were not detected in the cohort at large; however, these analyses were designed to look at average effects. The overall improvement in treadmill test performance for this cohort was relatively modest (median change in PWT =5.9 [IQR 1.7-10.2]), but the range of improvement was large (min=2.0 min, max=35 min), suggesting there may be individual level effects of SET training on the metabolome not yet uncovered in our previous analyses. We hypothesized that metabolite levels, static or dynamic, associate with an individual’s response to SET.

To test this hypothesis, we first calculated the individual response to SET. We modeled PWT on the treadmill test after SET based on peak walking time prior to SET using a generalized linear model. In this model, the slope (*β*) of the regression line represents the average effects of SET (Figure 4). To determine individual performance, we calculated the studentized residuals that capture the difference between the predicted and actual PWT on the post-SET treadmill test for each individual. We then tested the participant characteristics for an association with the studentized residual, or the standardized measure of individual improvement (SMII) in PWT, to identify confounding clinical factors that contributed to the subject-level variation effects of SET (Supplemental Table 15). Although age was nominally associated with the SMII in PWT (β=-0.05 per year, 95% CI [-0.1 - -0.01], p=0.02, FDR = 0.2), no demographic factors demonstrated an experiment wide significant association with the studentized residual, including BMI and T2D.

**Figure 4.**
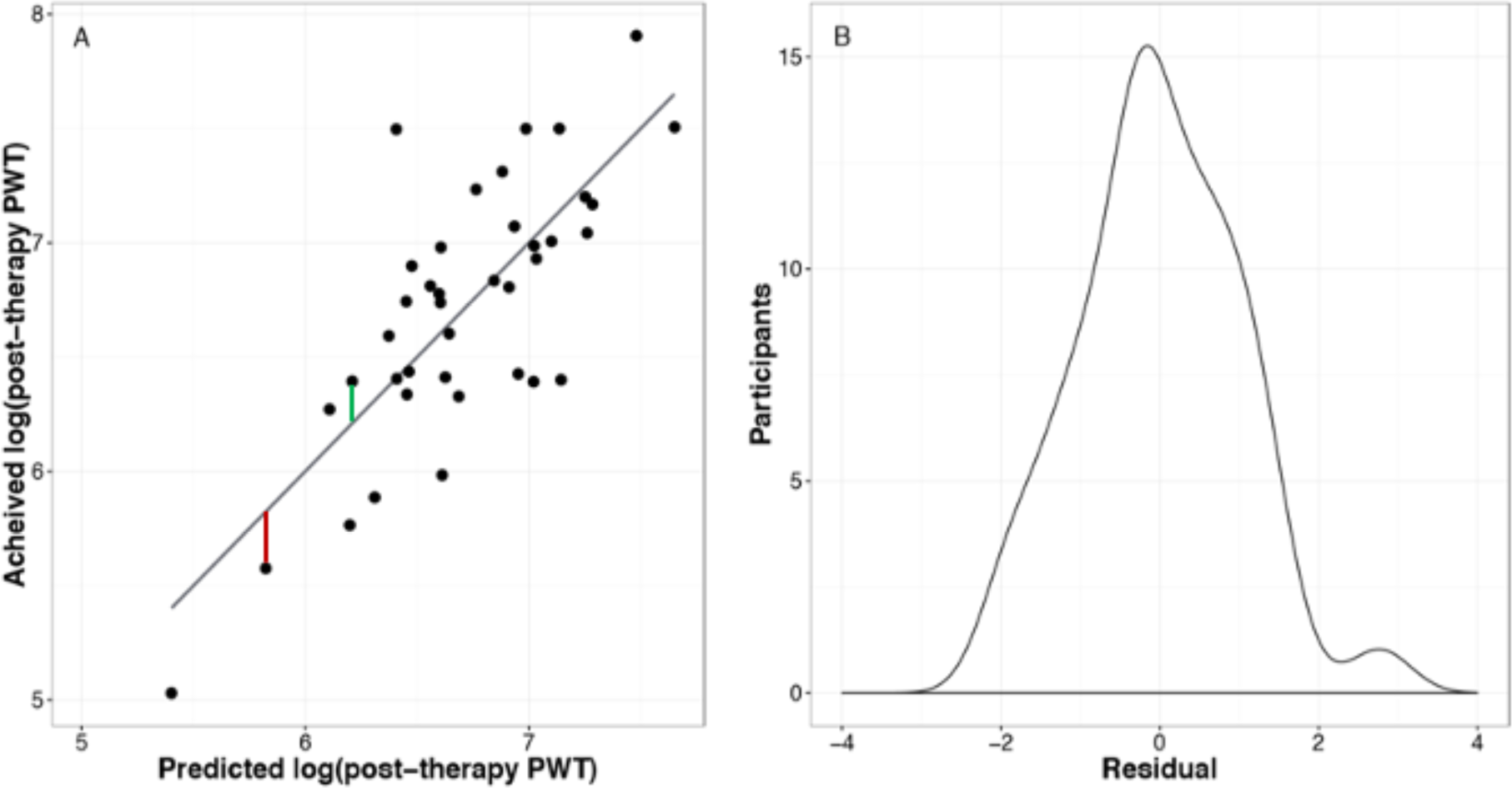
Adjusted individual subject-level variation in the effect of exercise training on Peak Walking Time (PWT). (A) A multivariable linear model was used to predict post-therapy log(PWT) based on pre-therapy log(PWT) and clinical factors. The slope (*β*) of the regression line represents the average effect of training on the change in log(PWT) across the cohort. Dots above the regression line represent subjects who performed better than predicted by the model and a positive distance was extracted as indicated by the green line. Dots below the regression line represent subjects who performed worse than predicted by the model and a negative distance was extracted as indicated by the red line. (B) A distribution of the studentized residuals, which represent the individual subject-level variation in the effects of exercise therapy that is not accounted for by the model.

Using the individual measure of improvement attributed to SET, we tested the association of baseline metabolite levels from the first study visit, with individuals measures of walking improvement using again the 5 different modeling approaches. Nominal significance in one model was achieved by 67 metabolites, nominal significance in two or more models was achieved by 32 metabolites, and none of the metabolites achieved experiment wide significance in one or more models, resulting in a total of 32 metabolites of significant interest (Supplemental Table 16). Homogeneous non-metal compounds (p = 0.02, FDR = 0.07), sphingolipids (p = 0.046, FDR = 0.07), fatty amides (p = 0.04, FDR = 0.1), and sterol and prenol lipids (p = 0.03, FDR = 0.07) were all nominally enriched metabolite classes (Supplemental Table 17, Supplemental Figure 7).

Higher levels of the homogeneous non-metal compound hydroxylamine were associated with diminished improvements in the SMII in PWT (Supplemental Figure 7). Interestingly, hydroxylamine is known to be utilized by fibroblasts to increase the surface expression of GLUT-1 transporters (53) and by skeletal muscle to increase glucose uptake through insulin receptor activation (53, 54). In PAD patients with claudication, PET imaging of skeletal muscle glucose uptake demonstrated calf muscle insulin resistance(55). Therefore, increased hydroxylamine in this study may reflect calf muscle insulin resistance and a mechanism for exercise limitation. The fatty amides metabolite class was positively associated with improvement in the SMII in PWT, meaning that higher levels of fatty amides before any exercise challenge were associated with larger improvements in the PWT from the first to the second treadmill test (Supplemental Figure 7).

We then examined whether metabolite changes induced by acute exercise prior to 12 weeks of SET was associated with improvement in walking following SET. We did this by modeling the association of changes in exercise before and after the first treadmill test with individual walking improvement. Nominal significance was achieved by 96 metabolites in one model, nominal significance in two or more models was achieved by 28 metabolites, and no metabolite achieved experiment wide significance in one or more models, resulting in a total of 28 metabolites of significant interest (Supplemental Table 18). These dynamic changes in metabolite classes uncovered the same metabolite classes found with resting plasma metabolites: homogeneous non-metal compounds (FDR = 0.02), sterol and prenol lipids (FDR = 0.046), and fatty amides (FDR < 1E-10) were all again experiment wide enriched and negatively associated with the SMII in PWT (Supplemental Table 19, Supplemental Figure 8). Therefore, large changes in these metabolite levels over the treadmill test prior to SET were associated with diminished response to SET.

### Changes in baseline metabolite levels in individuals who underwent SET uncover possible etiologies for large improvements in standardized measure of individual improvement in PWT

The pathophysiology underlying improvement in PWT among patients with PAD is currently incompletely understood. Characterizing changes in metabolite levels over the course of SET could provide insight into signaling or mechanistic pathways relevant to symptom relief or improvement in PAD. However, individual response to SET can vary widely and therefore metabolite changes may be inconsistent when observing average effects. To better understand mechanisms of improvement after SET on an individual level, changes in baseline metabolite levels were tested against the SMII in PWT.

To explore the impact of SET on metabolite levels, we first tested the association of the difference in baseline metabolite levels drawn before each treadmill test. We did this by comparing how changes in baseline metabolites (before any treadmill test) between samples collected before and after 12-weeks of SET associated with standardized measure of individual improvement in PWT from our regression model. Nominal significance in one model was achieved by 119 metabolites, nominal significance in two or more models was achieved by 59 metabolites, and experiment wide significance in one or more models was achieved by 14 metabolites, resulting in a total of 73 metabolites of significant interest (Supplemental Table 20). Fatty acyls (p = 0.045, FDR = 0.12) and homogeneous non-metal compounds (p = 0.04, FDR = 0.16 were nominally enriched in this model (Supplemental Table 21, Supplemental Figure 9). Organonitrogen compounds (FDR = 0.01) and fatty amides (FDR – 5E-03) were enriched at an experiment wide level and negatively associated with performance on the first treadmill test, meaning large resting increases in these metabolites after SET were associated with diminished response to SET (Supplemental Figure 9). The organonitrogen metabolite AEA and fatty amide metabolites interact with cannabinoid receptors (CB) (56, 57) and stimulate fast-muscle oxidative phenotypes that could impact exercise training (58).

### Dynamic changes in metabolite levels associate with individual improvement in walking performance

It is also possible that SET affects individual walking performance by changing the dynamic response of the metabolome to acute exercise. To explore this, the change in the metabolomic response to acute exercise before and after SET was tested for an association with the SMII in PWT. Nominal significance in one model was achieved by 89 metabolites, nominal significance in two or more models was achieved by 20 metabolites, and experiment wide significance in one or more models was achieved by 5 metabolites, resulting in a total of 25 metabolites of significant interest (Supplemental Table 22). Carbohydrates (FDR = 0.03), glycerolipids (FDR = 0.01), and eicosanoids (FDR < 1E=10) were experiment wide enriched in this model (Supplemental Table 23).

The eicosanoid class metabolite 5-KETE was positively associated with improvement after SET (Supplemental Figure 10), meaning larger increases in the levels of 5-KETE during acute exercise after SET, as compared to before SET, were associated with greater improvement in the SMII in PWT as a result of SET. In other words, larger increases in 5-KETE levels during an acute exercise challenge following SET is associated with greater improvement in PWT after SET. 5-KETE is a long-chain fatty acid, and these have been shown to be increased in patients with claudication compared to healthy patients (33).

## DISCUSSION

Metabolomics has been previously performed on patients with PAD to identify metabolites associated with disease progression (33, 34) as well as on a cohorts of healthy and insulin resistant people after exercise (41, 42, 59). However, metabolomics has not been used to examine the metabolic response to exercise training in patients with PAD. Our analysis of repeated measures generalized linear models suggested that on average, SET minimally effected the metabolome. Therefore, we sought to understand metabolic changes at specific time points instead of across all time points (Figure 1). To do this, we used five different models (GLM, LASSO, elastic net, random forest, SVR) to identify metabolites or changes in metabolites at specific time points that associated with either treadmill test performance or inter-individual variability in functional performance after SET. To further understand how classes of metabolites associate with exercise, enrichment analysis was performed (Figure 3). Our analysis of metabolite changes during SET identified the importance of analyzing inter-individual effects: some metabolite associations only became relevant when considering individual quantitative effects, not an average across the cohort. We verified previously known associations between SET and acylcarnitine as well as carbohydrates. Importantly, we found two pathways in skeletal muscle of relevance to individual response to SET which warrant further study: Anandamide (AEA) signaling and Arachidonic acid (AA) synthesis.

### Acylcarnitine depletion after exercise

Our results replicated known associations between acylcarnitine and improvement in walking time with SET (60). Circulating and intra-muscular levels of oxidative phosphorylation intermediates, including acylcarnitines, are known to be higher in patients with PAD (61). Low acylcarnitine levels could indicate this metabolite is being consumed by fatty acid catabolic pathways and therefore reflects high levels of fatty acid catabolism (62). This is likely reflective of mitochondrial dysfunction, which has been implicated in impaired skeletal muscle metabolism in PAD. Impaired mitochondrial function limits oxygen utilization and causes endothelial dysfunction, as well as muscle fiber disarray (61, 63, 64). Restoration of carnitine metabolism is associated with improvement in treadmill walking time (30, 65). In a prospective randomized interventional study, all participants had decreased acylcarnitine plasma levels after a 10 week exercise program (60). We have replicated those results in this study, where medium and long chain acylcarnitine levels are decreased after participants undergo SET, which suggest that SET augments mitochondrial function.

### AEA changes are associated with exercise tolerance

AEA is an organonitrogen compound derived from non-oxidative metabolism (66) that functions as an endocannabinoid signaling molecule. AEA is biosynthesized from its precursor glycerol-phospho-AEA via the enzyme glycerophosphodiesterase 1, resulting in glycerol-3-phosphate and AEA (67). In skeletal muscle, AEA exerts its effect on muscle metabolism through the membrane-bound G-proteins CB1 and 2 leading to changes in insulin sensitivity, different patterns of glucose uptake (68) and inhibition of myotube formation (58, 69) (Figure 5).

**Figure 5.**
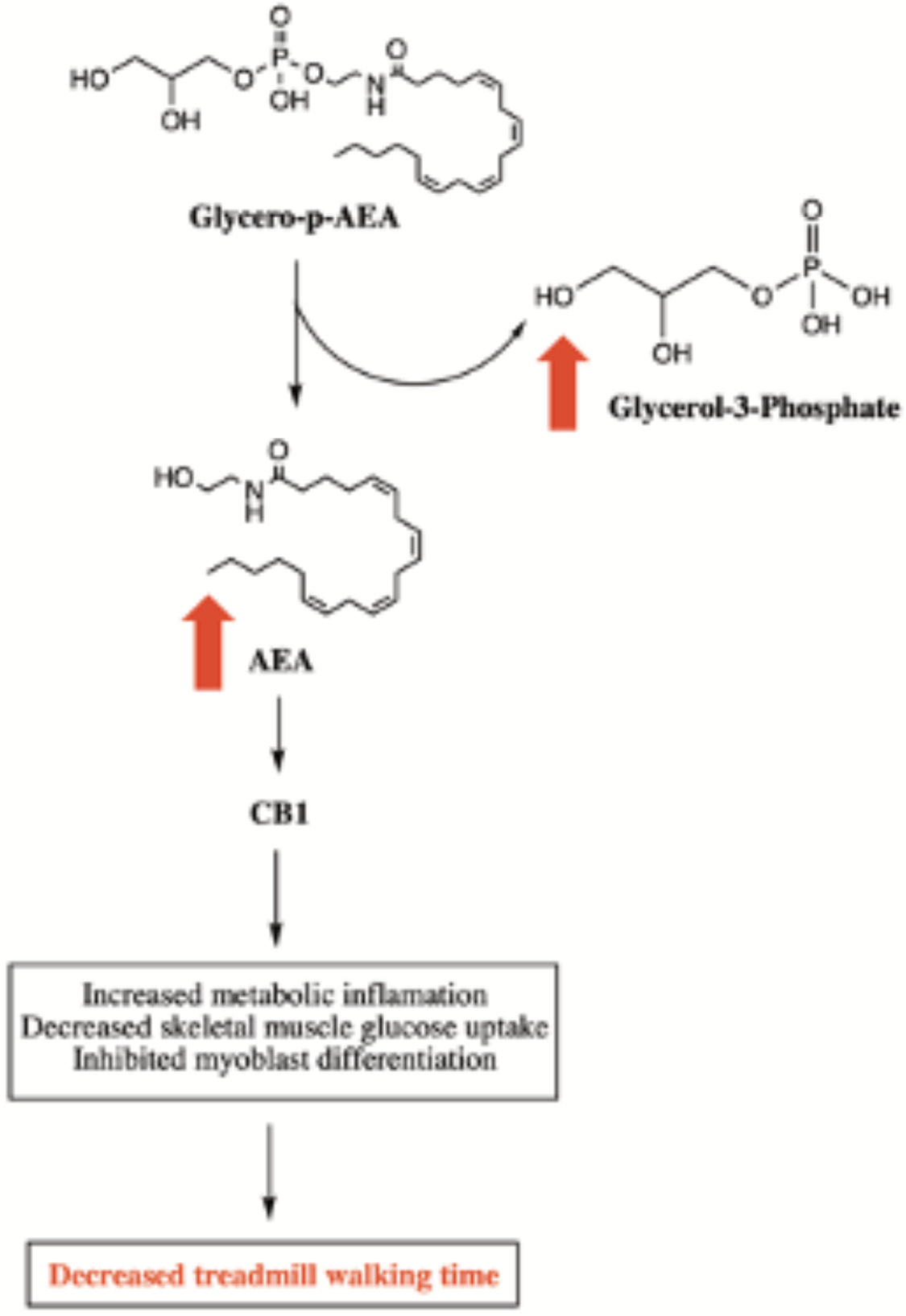
Synthesis and metabolism of AEA, an organonitrogen compound known to stimulate endocannanaboid receptors. Red arrows indicate that increased levels of metabolites through this pathway are associated with shorter walking times. Abbreviations: AEA, anandamide or N-arachidonoylethanolamine; CB1, cannabinoid receptor 1.

Although AEA is generally considered anti-inflammatory, AEA signaling through CB1 receptors is reportedly pro-inflammatory (70). Changes in circulating AEA and other AcylEAs during exercise have previously been described in healthy participants (71, 72), where the plasma concentration of AEA following exercise was shown to correlate with endocannabinoid activity. Therefore the effects of exercise on AEA levels can be directly linked to endocannabinoid signaling (73). Downstream activity of AEA signaling via the CB1 receptor has been shown to inhibit myoblast differentiation, expand the number of satellite cells, and stimulate the fast-muscle oxidative phenotype (74). Increased oxidation of fatty acids, but not glucose, can then lead to alterations in energy expenditure of these myoblasts (56, 58, 75). Chronic CB1 receptor stimulation has also been shown to increase metabolic inflammation in mice by inducing glucose intolerance (76). Even in acute instances, increased CB1 activation impairs skeletal muscle insulin sensitivity, which decreases glucose uptake in skeletal muscle (56, 77) and may contribute to an acquired skeletal muscle myopathy (78, 79). This is consistent with our analysis, which shows that increased plasma glucose levels over the course of acute exercise are associated with shorter or worse walking times. Additionally, studies have implicated a role of endocannanaboid signaling and increased intramuscular lipid oxidation (79) in pain and muscle soreness after exercise (80, 81), which could also be related to lipotoxic muscle pain.

In the current study, we found that high baseline levels of AEA were associated with a shorter peak walking time (Figure 5) in individuals with claudication. When observing individual level effects, we also found that participants that had large increases in baseline AEA levels and glycerol-3-phosphate levels after SET did not respond as well to SET. We hypothesize that increased AEA synthesis leads to increased skeletal muscle endocannabinoid signaling, decreased energy availability, more pain, increased walking dysfunction, and decreased response to SET (Figure 6). Additional studies are needed to link skeletal muscle insulin resistance to functional parameters in patients with PAD.

**Figure 6.**
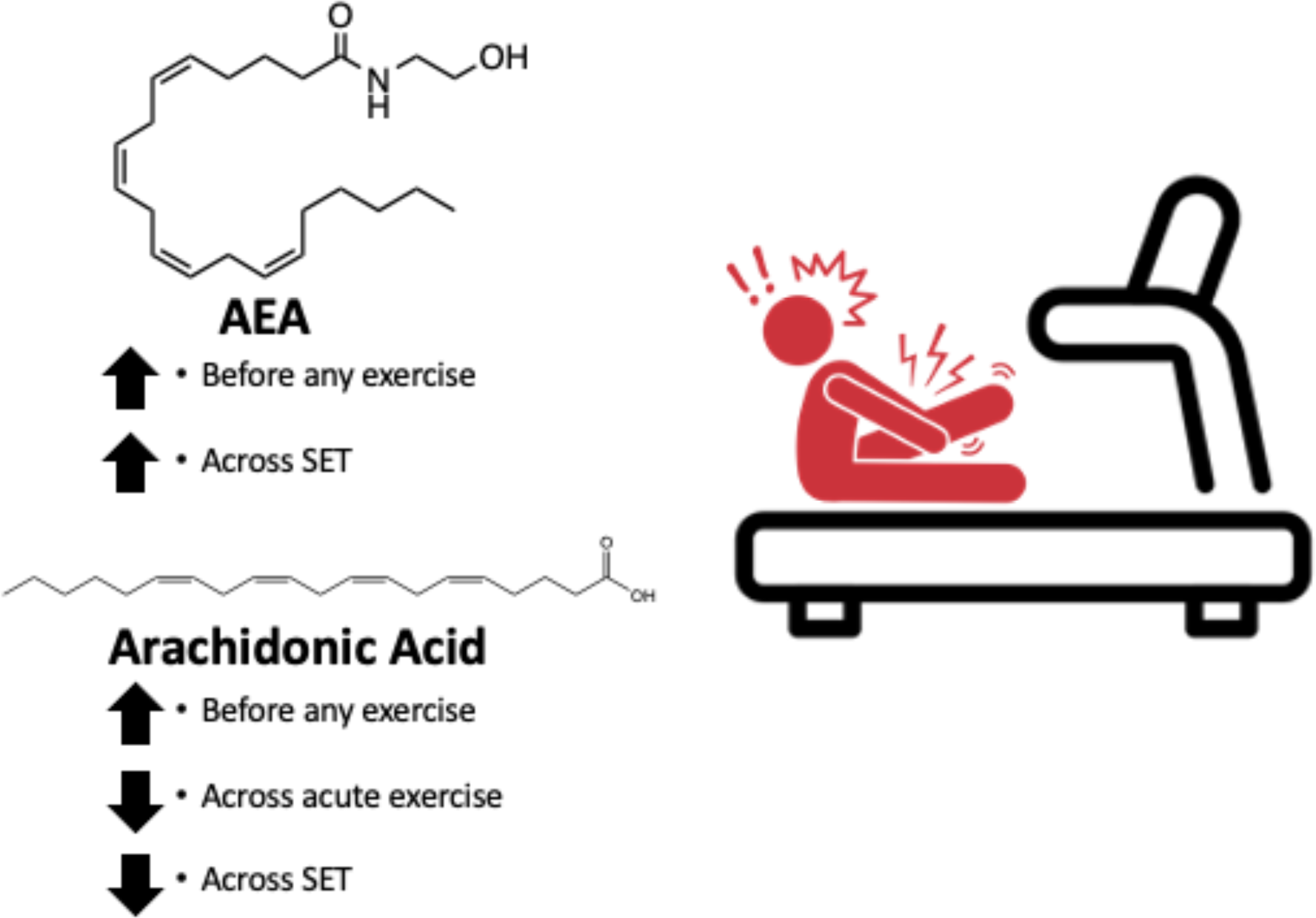
Anandamide (AEA) and arachidonic acid (AA) levels that associate with decreased walking time. Specifically, increased AEA before any exercise and a large increase in AEA after SET were associated with longer walking times. Increased AA before any exercise, small AA increase over acute exercise, and small AA increase after SET were associated with longer walking times.

### Arachidonic acid and chronic inflammation

The synthesis of AA is another key pathway that could contribute to the inflammatory response after exercise. AA is a polyunsaturated fatty acid present in phospholipids throughout the body, but is especially concentrated in skeletal muscle (82, 83). AA generally has pro-inflammatory and pro-aggregation effects (84). In this study, we found that increased AA at baseline was associated with a shorter walking time (Figure 7). Despite this, the participants that had the longest walking times, either before or after SET, had the largest increases in AA during the treadmill test. When analyzing the individual level effects of SET, we observed that large increases in baseline AA as a result of SET were associated with above average response to SET.

**Figure 7.**
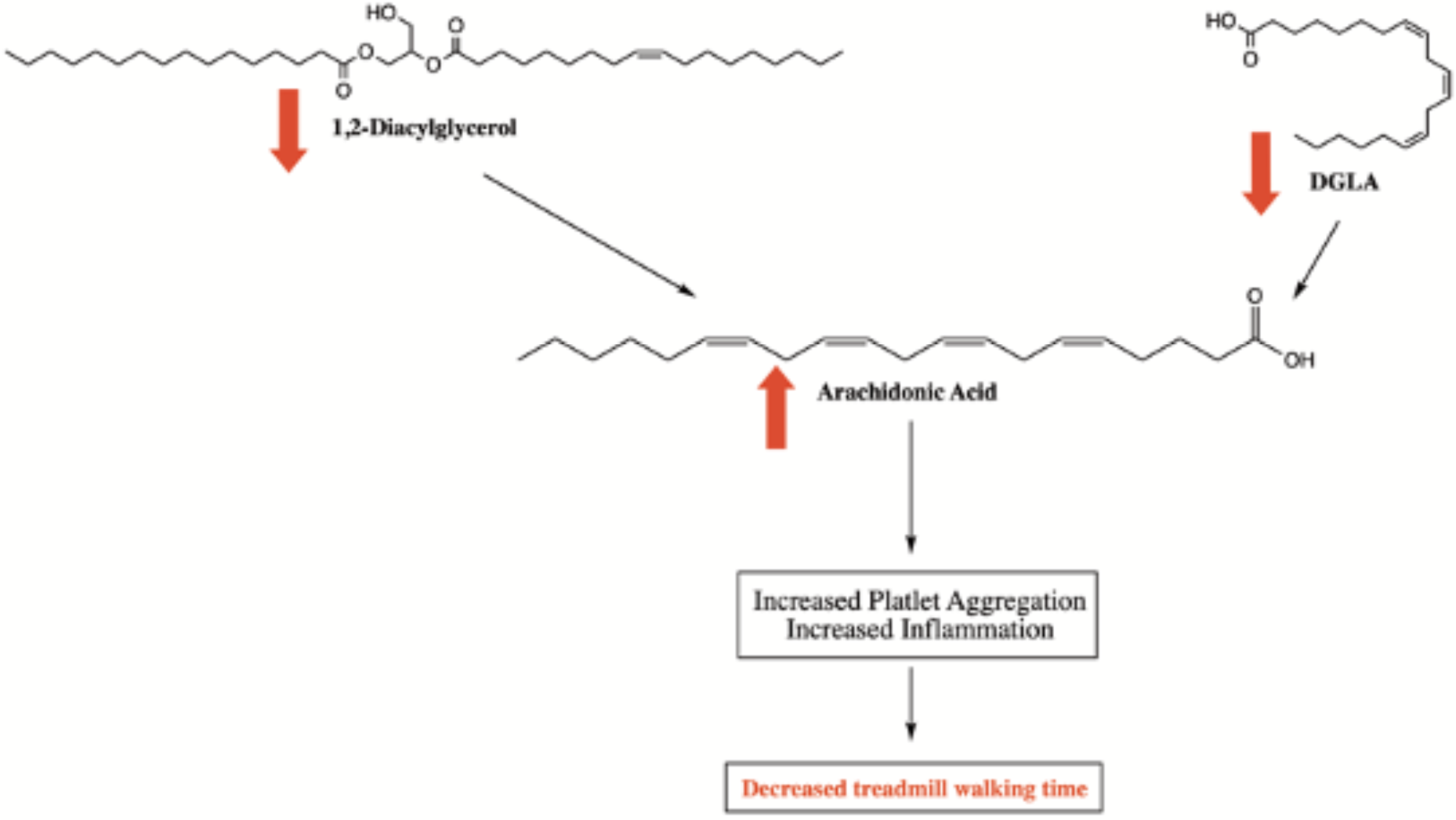
Pathways of arachidonic acid synthesis, which ultimately leads to inflammation. The direction of the arrows indicate the levels of the metabolite through this pathway. The color red indicates these metabolite levels are associated with shorter walking times. Abbreviations: DGLA, Dihomo-γ-linolenic acid.

AA and its downstream metabolites are known to play roles in both the initiation and resolution of inflammation of several disease states, including obesity, diabetes, NAFLD, and cardiovascular disease (44). The metabolites of AA are generally inflammatory: while cytochrome P450-dependent epoxygenated AA metabolites are largely anti-inflammatory, the rapid breakdown of these again yields pro-inflammatory signals (85). AA derivatives from the lipooxygenase pathways are involved in promoting hyperalgesia and derivatives from the cyclooxygenase pathway are involved in the production of thromboxane A2 and prostaglandins (86), which contribute to an inflammatory response.

Based on our findings and known AA biology, we hypothesize that high levels of AA before exercise are pro-inflammatory and lead to decreased walking ability in individuals with claudication. AA is produced during exercise and individuals with longer walking times are able to tolerate more AA production. SET helps to train individuals with claudication to withstand higher levels of AA and inflammatory signaling, resulting in longer walking times (Figure 6). In short, we hypothesize if patients can tolerate this inflammation or pain longer, they may be able to walk further.

Our hypothesis is supported by the measurements of AA precursors (Figure 7). AA can be synthesized by many pathways, including from the precursor dihomo-γ-linolenic acid (DGLA) via the enzyme Δ5 desaturase (87). Diacylglycerol (DAG) can also be broken down via diacylglycerol lipase into 2-arachidonoylglycerol, which is further broken down via monoacylglycerol lipase resulting in AA production (88). We found that increased baseline levels of plasma DGLA are associated with longer walking times. Elevations in DGLA are known to correlate with decreased levels of AA providing corollary support to our hypothesis (89). Our analyses also demonstrated that increasing concentrations of DAG after treadmill testing were associated with longer walking times. Taken together, this evidence supports the idea that longer walking times overall are associated with increased levels of AA precursors DAG and DGLA, and decreased levels of AA itself.

## Conclusions

Although SET minimally effected the metabolome on average, we found interesting metabolic patterns when analyzing inter-individual effects (Figure 3). Increased levels of plasma carbohydrates during acute exercise may trigger increased IL6 signaling, citrate synthase activity, and intramuscular lipid oxidation, which all contribute to muscle pain (45, 79). We also found low acylcarnitine levels after participants underwent SET, which may indicate consumption of carnitine by fatty acid catabolic pathways and restoration of carnitine metabolism (62). Importantly, we also found two pathways in skeletal muscle of relevance to individual response to SET which warrant further study: high levels of AEA are linked to CB1 signaling and activation of inflammatory pathways (73). This alters energy expenditure in myoblasts (56, 58, 75) by decreasing glucose uptake (56, 68) and may induce an acquired skeletal muscle myopathy (78, 79). The second pathway that contributes the inflammatory response after exercise is AA synthesis (84, 85). SET may help participants tolerate increased levels of AA and inflammation produced during exercise, resulting in longer walking times.

## Limitations

There are limitations of this study concerning both experimental design and metabolite measurements. This study was a secondary analysis of blood samples drawn at time points not designed to optimize sample integrity for metabolomics: non-fasting blood samples were used for analysis and this work did not include participant diet records to control for the impact of diet on the metabolome (90). Furthermore, anti-inflammatory medication use was not recorded. Ibuprofen is a common medication known to directly affect the concentration of several metabolites measured in this study (91). Lack of control for diet and medications in the experimental design would have introduced noise and thus would be expected to bias our results to the null. Although this study did not have a control group, we have considered each participant’s baseline metabolite levels in our analysis. In terms of metabolite levels, some metabolites measured are involved in multiple pathways and have multiple breakdown products with opposing effects. For example, metabolites of AA can have both pro and anti-inflammatory effects, which makes interpretation of these results more challenging (92). Future work is needed to better validate our metabolic patterns found in a larger participant cohort with an experimental design optimized specifically for metabolomic analysis, including fasting blood draws, monitored diet status, and records of medication intake. Future work could also include targeted pathway analysis to uncover interactions between metabolites of significant interest found in this study: AEA, AA, isothreonic acid, and acyl carnitines in relationship to free fatty acid levels.

## METHODS

The study was reviewed by the University of Pennsylvania Institutional Review Board for Human Subjects Research. All participants provided written informed consent prior to inclusion in this study: NCTID 2RO1HL075649-06A2.

### Randomized Control Trial Design

130 individuals with peripheral artery disease, as defined by ankle brachial indices ≥ 0.4 and ≤ 0.8 and the presence of classic claudication symptoms, were randomized to 12 weeks of a standard, validated SET program (93) or usual medical care. Both prior to initiating and on completion of the exercise program, participants underwent the Gardner graded treadmill walking test (37) to determine peak walking time (PWT) (94). Venous blood was sampled before and after each treadmill test using sodium citrate collection tubes, and platelet free plasma was stored at -80°C. The current analysis was restricted to individuals who were randomized to the exercise program arm, completed at least 80% of prescribed sessions, and had all 4 plasma samples available. 43 participants in the study met these criteria. Based on available resources, the 40 participants who completed the greatest number of training sessions were selected for metabolite analysis.

### Measurement of Metabolites

The plasma samples of the 40 participants were analyzed using one targeted and two untargeted metabolite panels: primary metabolites were measured by GC-TOF, complex lipids were measured by LC-TOF were measured by UPLC QTRAP. Lipid mediators including oxylipins, endocannabinoids, and fatty acids were isolated from plasma using Ostro Sample Preparation Plates (Waters; Milford, MA) in the presence of BHT/EDTA and quantified by UPLC-MS/MS using internal standard methods. Briefly, in the Ostro Plate well, 100µl plasma aliquots were mixed with 5µl of BHT/EDTA (1:1 v/v MeOH/water) and 5µl methanol containing a suite of deuterated surrogates. The samples were then mixed with 300µL of acetonitrile with 1% formic acid and eluted with vacuum into 200µL glass inserts containing 10µl of 20% glycerol in methanol. Solvent was removed by vacuum centrifugation and the glycerol plug was stored at -80C° until analysis. Prior to acquisition, samples were reconstituted in 100µl of 1:1 methanol:acetonitrile containing 100nM of 1-cyclohexyl ureido, 3-dodecanoic acid (CUDA) and 1-phenyl ureido, 3-hexadecanoic acid (PUHA). Residues were separated on a 2.1 x 150mm 0.17µm BEH column (Waters) and detected by electrospray ionization with multi reaction monitoring on a API 6500 QTRAP (Sciex; Redwood City, CA) and quantified against 7-9 point calibration curves of authentic standards using modifications of previously reported methods (42).

### Data Processing

Prior to association testing, metabolite levels were natural log transformed and pareto scaled. Metabolites with any incomplete data across samples were removed from this analysis. Only peaks corresponding to known metabolites were considered in the analysis, for a total of 122 known metabolites in the primary metabolite panel, 306 metabolites in the complex lipids panel, and 64 metabolites in the lipid mediators panel (Supplemental Figure 1).

### Studentized Residual to Model Subject-Level Variation

To model individual variation in the response to SET, a generalized linear model was constructed that predicted the log-transformed PWT [log(PWT)] measured on the post-training treadmill test based solely on log(PWT) from the pre-training treadmill test. The slope (*β*) for the pre-training log(PWT) represents the average effect of training on the change in log(PWT) across the cohort, with individual variation represented by the studentized residuals extracted from the model.

### Metabolite Annotation

Metabolites were annotated with a super class, main class and sub class by compound name using the RefMet database from the UCSD Metabolomics Workbench (38). The most specific classification that yielded groups of four or more metabolites in each dataset was used for hypergeometric enrichment. Super class designations were used for the primary and the complex lipids datasets, main class designation was used for the lipid mediators dataset.

### Statistical Analysis

All data handling and analysis were performed using R v3.6.2.

### General Linear Models

General linear models were fit using the base glm function in R v3.6.2 to test for the association of various phenotypes as exposures with individual baseline metabolite levels as the outcome. Unless otherwise specified, these baseline models included age and sex as covariates. Repeated measures general linear models were fit using the ‘lmerTest’ (95) package in R to test the association between individual metabolite levels and various outcomes. Models that tested the association of individual metabolite levels with walking time based outcomes utilized post-hoc contrast statements and the ‘multcomp’ (96) package in R to model this association at various timepoints.

### Machine Learning Approaches

We sought to understand metabolic changes at specific time points using five different models: GLM, LASSO, elastic net, random forest, SVR (Figure 4). The four machine learning models used are described below.

RF models were generated using the package ‘rfPermute’ in R (97). RF is an is an extension of bagged decision trees that allows for the assessment of a predictor’s importance in modeling the outcome (98, 99). 2000 trees were grown and permuted 50 times. Increase in the mean squared error of the model (estimated with an out-of-bag cross validation) as a result of an individual predictor being permuted and its associated probability value were extracted from each RF model.

Least absolute shrinkage and selection operator (LASSO) and elastic net regression were performed using the package ‘GLMNET’ in R (100). LASSO and elastic net regression are extensions of generalized regression that minimize residual sum of squares by shrinking some coefficients to zero using penalty terms *L_1_-*norm and *L_1_-*norm + *L_2_-*norm respectively, thereby eliminating them from the model. This allows for stable model selection and regression without over-fitting (101, 102). A 10 fold cross validation of both a LASSO and an elastic net regression were performed separately to produce an optimal tuning parameter (minimum value of lambda) that would minimize the cross-validated error and also protect against tests already run by the model. Metabolites with coefficients not equal to zero were considered to have a nominally significant association with the outcome.

Support vector regressions are an adapted form of support vector machines used when the dependent variable is continuous rather than categorical (103). A 10 fold cross validation of the support vector regression was performed and weights, defined as the dot product of the mapped vectors and the support vectors of the regression, were extracted for each predictor (104). Support vector regression was performed using the ‘e1071’ package in R (105). In practice, using LASSO and elastic net regression for association testing translates to classifying the excluded variables as unimportant and the included predictors as important. Furthermore, support vector regression for association testing neglects measuring error for each individual weight. To determine a probability value for LASSO and elastic net regression coefficients and support vector regression weights, each regression was bootstrapped 1000 times to estimate the standard error (SE) of each coefficient/weight (106, 107). The SEs were then used to generate a probability value based on a normal distribution.

### Significant Thresholds

Analyses testing the association of various phenotypes with baseline metabolite levels and the association of metabolite levels with average responses to treadmill testing and SET defined experiment wide significance as a Benjamini-Hochberg FDR of < 0.05 as calculated individually for each analyzed panel of metabolites.

In the analyses investigating metabolite levels as predictors of peak walking time and individual improvement, predictors were defined as all metabolites or their difference in the above described panels. The outcome was defined as either pre-therapy log(PWT) or the above described studentized residual. Five different models were used to incorporate the advantages from each model described below and increase detection of true positives: GLM, LASSO, elastic net, random forest, support vector machines (Figure 4). To limit detection of false positives, metabolites of significant interest were defined as being nominally significant on 2 or more models or experiment wide significant on one model. For all tests, nominal significance was defined as p < 0.05 and experiment wide significance was defined as a Benjamini-Hochberg FDR of < 0.05 for the number of known metabolites in the analyzed panel.

### Hypergeometric Enrichment

Hypergeometric enrichment analysis was performed using the ‘phyper’ function in R among the for the RefMet annotated classes for metabolites that passed the previously described significance thresholds for each experiment and each dataset individually. Hypergeometric enrichment analysis uses a hypergeometric distribution to compute the probability of seeing more observations of a certain category (RefMet annotated class) in *n* draws without replacement. The hypergeometric distribution is parameterized by three quantities, the total number of metabolites, the number of metabolites of interest and the number of metabolites in the group of interest. Nominal significance was defined as p < 0.05. Multiple testing correction was done using a Benjamini-Hochberg FDR for all metabolite groups tested in each dataset and experiment separately. Experiment wide significance was defined as FDR < 0.05.

## SUPPLEMENTAL INFORMATION

Data and code that were used in this study are available on request from the corresponding author (S.M.D.).

## AUTHOR CONTRIBUTIONS

Designing research studies: SMD, TRB, NLT. Acquiring data: SMD, TRB, NLT. Analyzing data:TRB, NLT, SMD, KB, GS. Writing and crucial editing: SMD, JWN, TRB, NLT, HJC, KB, GS, RJ, SR, OF, TFF, FWW.

## Data Availability

Additional data that were used to generate the Figures in this study are available on request from the corresponding author (S.M.D.)

## ACKNOWLEDGEMENTS

We would like to acknowledge Dr. Emile R. Mohler, now deceased, whose dedicated work to understanding vascular health provided the initiative and basis for this research study. This research supported by NIH NCATS UL1TR000003. S.M.D was also supported by funding from the Department of Veterans Affairs IK2-CD001780. The authors wish to thank patient participants; without their assistance, this research would not have been possible. This publication does not represent the views of the Department of Veterans Affairs, the US Food and Drug Administration, or the US Government.

